# Education as a Proxy for Cognitive Reserve: Moderating Effects on White Matter Hyperintensity Burden in Healthy Aging and Cognitive Decline

**DOI:** 10.1101/2024.09.15.24313717

**Authors:** Odelia Elkana, Iman Beheshti, Alzheimer’s Disease Neuroimaging Initiative

**Affiliations:** Behavioral Sciences, Academic College of Tel Aviv-Yaffo, Tel Aviv, Israel; Department of Human Anatomy and Cell Science, University of Manitoba, Winnipeg, MB, Canada

**Keywords:** Cognitive Reserve, Education, White Matter Hyperintensities, Cognitive Decline, Mild Cognitive Impairment, Aging, Neuroimaging

## Abstract

**Background:** Cognitive reserve, often approximated by levels of education, is thought to protect against the deleterious effects of brain pathology on cognitive function. White matter hyperintensities (WMHs) are commonly associated with aging and cognitive decline, and higher WMH burden has been linked to the progression from healthy cognitive status (HC) to mild cognitive impairment (MCI). Understanding how cognitive reserve, as indicated by education, influences the relationship between WMH burden and cognitive outcomes can provide valuable insights for interventions aimed at delaying cognitive decline.

**Objective:** This study investigates the moderating role of education, as a proxy for cognitive reserve, on the relationship between WMH burden and the transition from HC to MCI.

**Methods:** Data were obtained from the Alzheimer’s Disease Neuroimaging Initiative (ADNI) database, focusing on participants classified as cognitively healthy at baseline. A total of 153 cognitively healthy adults at the baseline were split into two groups: one group (n=85) remained cognitively healthy for at least 7 years, while the other group (n=68) progressed to MCI within 7 years. A multiple linear regression model was used to examine the interaction between group membership, baseline age, education, and sex in predicting WMH loads. The primary focus was on the interaction between group membership and education to assess the protective effect of cognitive reserve.

**Results:** The regression model explained 18.5% of the variance in WMH load. The analysis revealed statistically significant interaction between group membership and education on WMH loads (Interaction term: β = -0.097, p = 0.047), indicating that higher education levels are associated with a reduced WMH burden among individuals who progressed to MCI. The main effect of education alone was not significant, nor were the interactions involving sex (p > 0.05).

**Conclusion:** These findings support the hypothesis that education, as a proxy for cognitive reserve, provides a protective effect against the accumulation of WMH burden in older adults. The results suggest that higher cognitive reserve may mitigate the impact of neurodegenerative processes, thereby delaying the transition from HC to MCI. This underscores the importance of educational attainment in the preservation of cognitive health during aging.

## 1- Introduction

Cognitive reserve (CR) refers to the brain’s ability to optimize or maximize performance through differential recruitment of brain networks or alternative cognitive strategies, thus allowing individuals to cope better with brain pathology (Stern, 2009). The concept of cognitive reserve emerged from observations that individuals with similar levels of brain pathology often exhibit different levels of cognitive function (Stern, 2002). Education is frequently used as a proxy for cognitive reserve, with higher levels of education associated with a greater capacity to withstand the effects of brain pathology, such as Alzheimer’s disease (AD) and other forms of dementia (Stern, 2012).

White matter hyperintensities (WMHs) are a common form of brain pathology observed on T2-weighted fluid-attenuated inversion recovery (FLAIR) MRI scans. These hyperintensities are considered markers of small vessel disease and are frequently observed in older adults, with their prevalence and burden increasing with age (Prins and Scheltens, 2015). WMHs have been linked to a variety of adverse outcomes, including cognitive decline, an increased risk of dementia, and even mortality (Debette and Markus, 2010; Gouw et al., 2008). The exact mechanisms through which WMHs contribute to cognitive decline are not fully understood, but they are believed to disrupt the integrity of white matter tracts, leading to impaired communication between brain regions (Vernooij et al., 2007).

The relationship between WMHs and cognitive function is complex and is thought to be moderated by factors such as cognitive reserve. Individuals with higher cognitive reserve, often inferred from educational attainment, may exhibit better cognitive function despite having a high burden of WMHs (Brickman et al., 2011). This protective effect of cognitive reserve is thought to be due to the brain’s ability to compensate for damage by utilizing more efficient cognitive networks or by engaging in alternative cognitive strategies (Stern, 2012). Research has shown that higher levels of education are associated with a reduced impact of WMHs on cognitive function. For instance, a study found that the negative impact of WMHs on cognitive decline was less pronounced in individuals with higher educational attainment (Dufouil et al., 2003). Similarly, it has been documented that the association between WMHs and cognitive impairment was weaker in individuals with higher levels of education, suggesting that cognitive reserve may mitigate the detrimental effects of WMH burden (Murray et al., 2012).

While previous studies have established a link between cognitive reserve and resilience to brain pathology, less is known about the specific role of education in moderating the relationship between WMH burden and the transition from cognitively healthy (HC) status to mild cognitive impairment (MCI). MCI is considered a transitional stage between normal aging and dementia, and individuals with MCI are at a higher risk of progressing to AD (Petersen et al., 2014). Understanding the factors that influence the progression from HC to MCI is critical for developing interventions aimed at delaying or preventing cognitive decline.

In this study, we aim to investigate the moderating effect of education on the relationship between WMH burden, and the risk of cognitive decline from HC to MCI. By examining the interaction between group membership (HC remain vs. HC to MCI) and education, we seek to elucidate whether higher educational attainment, as a proxy for cognitive reserve, can provide a protective effect against the accumulation of WMHs and the associated risk of cognitive decline. This research has the potential to inform strategies for enhancing cognitive reserve through educational and other cognitive enrichment activities, with the goal of promoting cognitive health in aging populations.

## 2. Material and methods

### 2.1. Participants

The data for this study was obtained from the Alzheimer’s Disease Neuroimaging Initiative (ADNI) database, with the dataset downloaded in July 2024. The ADNI database includes a variety of participants categorized by cognitive status, such as HC, MCI, and AD.

For this study, we examined participants classified as HC at baseline (N = 2,564) and assessed their cognitive status over a seven-year period. Based on their cognitive trajectories, participants were categorized into two groups:

- Group 1 (HC remain): This group comprised individuals who were cognitively healthy at baseline and remained cognitively stable throughout the seven-year follow-up (n = 151). WMH data were available for 85 participants in this group.
- Group 2 (HC to MCI): This group included participants who were cognitively healthy at baseline but transitioned to MCI during the seven-year follow-up (n = 121). WMH data were available for 68 participants in this group.

### 2.2. Definition of Cognitive Status

Cognitive status data for individuals was sourced from ADNI, which conducts thorough comprehensive clinical diagnostic evaluation including various cognitive assessments. These assessments have evolved over time. In the ADNI study, individuals with HC have a Clinical Dementia Rating (CDR) score of 0, indicating no significant cognitive impairment, and their Mini-mental State Exam (MMSE) scores typically range from 28 to 30. Conversely, those with MCI include a CDR global score of 0.5 and memory box score must be at least 0.5. Mini-mental State Exam (MMSE) and Wechsler Logical Memory II sub-scale are also used to establish a MCI diagnosis (with specific cut off scores based on educational level). For more detailed information on cognitive status assessment in ADNI, visit: https://adni.loni.usc.edu/data-samples/adni-data/study-cohort-information/

#### 2-2 Standard protocol approvals, registrations, and patient consents

All ADNI sites received approval from an ethical standards committee, and all participants provided written consent. More information about ADNI protocol approvals, registrations, and patient consents can be found at: https://adni.loni.usc.edu/.

#### 2-3 WMH measurements

WMH volumes were obtained from the ADNI dataset through high-resolution 3D T1 and FLAIR sequences. Briefly, each 3D T1 image was aligned to a common template atlas using non-linear methods, and non-brain structures were removed from the images. The FLAIR image was then aligned to the 3D T1 image using the FLIRT method from the FSL toolbox. WMH volume was estimated using a modified Bayesian probability structure. Additionally, the Total Intracranial Volume (TIV) was calculated from the 3D T1 images. To account for differences in head size, WMH volume was divided by TIV for each sample to obtain normalized WMH (nWMH) volume, which was then expressed as a percentage.

#### 2-4 Statistical Analysis

To investigate the moderating effect of education on the relationship between WMH burden and cognitive decline, a multiple linear regression analysis was conducted. The dependent variable was the nWMH, and the independent variables included group membership (HC remain vs. HC to MCI), education (years), and sex. The primary focus was on the interaction term between group membership and education (Group × Education) to assess whether education moderated the impact of WMH burden on the transition from HC to MCI. The model also included interaction terms between education and sex (Education × Sex) and group membership and sex (Group × Sex) to explore potential sex differences in the moderating effect of education.

The regression model was adjusted for baseline age, given its known impact on both WMH burden and cognitive function. A partial correlation was used to analyze the connection between WMH loads and education level, with adjustments made for baseline age and sex. Additionally, a Pearson correlation was employed to investigate the relationship between WMH loads and baseline age. All statistical analyses were performed using Python’s `statsmodels` library, with the significance level set at p < 0.05. P-values were adjusted using the false discovery rate (FDR) technique.

## 3. Results

### 3.1 Clinical and demographical characteristics

There were no statistically significant differences between the two groups in terms of demographic variables (i.e., age, education, sex) and cognitive symptoms (i.e., MMSE, CDR) (q > 0.05, FDR-corrected; see Table 1) at the baseline. Although the group 1 (HC remain) had a slightly lower WMH load than the group 2 (HC to MCI) at baseline, the difference was not statistically significant (t-test = 1.37, p = 0.17, independent student t-test). In the group 2, the progression from normal cognition to MCI varied between 6 months and 6 years after the baseline measurement, with a mean duration of 3.28 (±3.71) years.

**Table 1:** The baseline Clinical demographics of individuals included in this study.

| Characteristics | Group 1 | Group 2 |
| --- | --- | --- |
| **N (male %)** | 85<br>(51.76%) | 68<br>(57.35%) |
| **Demographics** | Mean<br>(SD) | Mean<br>(SD) |
| Age, years | 76.05<br>(5.92) | 77.05<br>(6.18) |
| Education, years | 16.67<br>(2.63) | 16.14<br>(2.63) |
| **Cognitive symptoms** |  |  |
| MMSE | 29.18<br>(1.14) | 28.72<br>(1.41) |
| CDR | 0.00<br>(0.00) | 0.00<br>(0.00) |
| **WMH** |  |  |
| %WMH | 0.46<br>(0.72) | 0.63<br>(0.83) |

**Note:** N: Number of subjects; ; WMH: White Matter Hyperintensity. Significance levels were assessed using a t-test for continuous variables and a Chi-square test for categorical variables, comparing the HC and MCI groups, with false discovery rate (FDR) correction applied for multiple comparisons (\*P < 0.05, \*\*P < 0.001, \*\*\*P < 0.0001). All individual at baseline, exhibited no indications of cognitive impairment or neurological disorders. The cognitive status is reported in accordance with the criteria established by the Alzheimer's Disease Neuroimaging Initiative (ADNI). Group 1 (HC remain) consists of individuals stable over seven years, while Group 2 (HC to MCI) includes those who progressed from healthy to MCI.

### 3-2 Modeling WMH load at baseline

The multiple linear regression model accounted for 18.5% of the variance in white matter hyperintensity (WMH) load, indicating that group membership, education, sex, and their interactions are important factors in predicting WMH burden, though additional factors likely contribute. A significant main effect of group membership on normalized WMH (nWMH) was observed, with the MCI group showing higher nWMH levels compared to the CH group (β = 1.812, p = 0.019). This finding highlights that individuals who transitioned from healthy cognition to MCI exhibited a greater WMH burden.

Education did not have a statistically significant main effect on WMH load (β = 0.0292, p = 0.436), indicating that education alone does not predict WMH burden without considering group membership. However, a significant interaction effect between education and group membership was identified (β = -0.0972, p = 0.047), suggesting that the protective effect of education against WMH accumulation is more pronounced in the MCI group. Specifically, higher education levels were associated with a lower WMH burden among individuals who transitioned to MCI, suggesting that cognitive reserve, as indicated by education, may mitigate the impact of neurodegenerative processes on WMH accumulation.

Further analysis of the relationship between WMH load and education within each group revealed a significant negative association in the MCI group (r = -0.26, p = 0.03, partial correlation adjusted for age and sex), indicating that higher education was linked to lower WMH burden in this group. In contrast, no significant association was observed in the CH group (r = 0.05, p = 0.53, partial correlation; Fig. 1a). The analysis of sex as a factor revealed that it did not have a statistically significant impact on WMH burden (β = 0.2354, p = 0.778), indicating that sex does not play a major role in the variation of WMH load within this model. Conversely, baseline age showed a strong and significant association with WMH load (β = 0.0375, p < 0.001), emphasizing the role of aging in WMH accumulation. Lastly, the relationship between WMH loads and age was consistent across both groups, with similar correlations observed in the CH group (r = 0.26, p = 0.012) and the MCI group (r = 0.26, p = 0.027; Pearson correlation; Fig. 1b).

**Figure 1:**
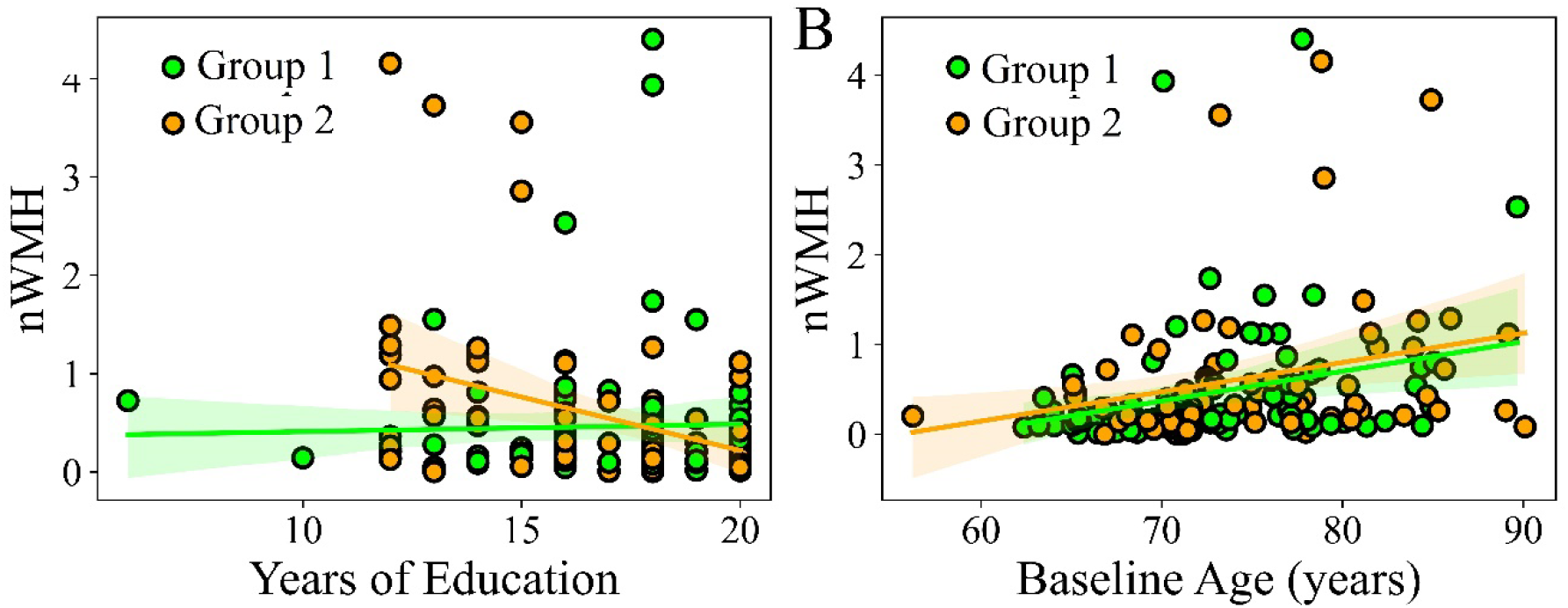
(A) Association between normalized WMH volume and education level within each group. (B) Association between normalized WMH volume and baseline age for each group. WMH volume is normalized by total brain volume. The relationship between WMH volume and education level was evaluated using a partial correlation test, adjusted for age and sex. The relationship between WMH volume and baseline age was also assessed using a partial correlation test. Group 1 (HC remain) consists of individuals stable over seven years, while Group 2 (HC to MCI) includes those who progressed from healthy to MCI.

**Table 1:** Summary of Multiple Linear Regression Analysis Predicting WMH burden including Model Fit Statistics and Regression Coefficients.

| Statistic | Value |
| --- | --- |
| R-Squared | 0.165 |
| Adjusted R-Squared | 0.125 |
| F-Statistic | 4.105 |
| P-Value Of F-Statistic | 0.000384 |

| Variable | B (Coefficient) | P-Value |
| --- | --- | --- |
| Intercept | -2.6775 | 0.010 |
| Group | 1.8123 | 0.019 |
| Sex | 0.2354 | 0.778 |
| Group × Sex | -0.2281 | 0.382 |
| Education | 0.0292 | 0.436 |
| Group × Education | -0.0972 | 0.047 |
| Education × Sex | -0.0225 | 0.645 |
| Age | 0.0375 | < 0.001 |

**Note:** Group=HC remain, HC to MCI; Sex=male, female

## 4- Discussion

In this study, we used a multiple linear regression model to examine how education and group membership interact to assess the potential protective effect of education against neurodegenerative processes. By considering education as a moderator between WMH burden and cognitive decline, rather than focusing solely on correlations, we offer a more nuanced understanding. Additionally, normalizing WMH volume as a percentage of total intracranial volume enhances the accuracy and reliability of our findingsproviding a clearer view of the protective effects of education.

As expected, increasing age was significantly associated with increasing WMH loads in both groups. This trend can be attributed to the fact that as individuals age, there is a decrease in cerebral blood flow and vascular density, which can result in ischemic damage in the brain’s white matter. This decline is often worsened by conditions like hypertension and atherosclerosis, which are more prevalent in older adults (Peters, 2006; Zhuang et al., 2018). Additionally, the rise in chronic inflammation as individuals age is associated with the formation of WMHs (Raz et al., 2012). Our research also considers sex as a variable, exploring potential sex differences in the moderating effect of education on WMH burden an area that has been underexplored in the literature. This inclusion adds another layer of complexity to our findings, helping to address a significant gap in understanding how cognitive reserve may differ between men and women. The statistical model we used did not find a significant main effect of sex on WMH burden at the baseline, indicating that sex does not have a notable impact on WMH burden. This result aligns with previous studies that have also reported minimal differences in WMH burden between cognitively healthy men and women (Morrison et al., 2024).

It is important to note that our study examined sex differences in the global burden of WMH. At the regional level, men may exhibit increased WMH progression in the occipital regions but show less progression in the frontal, total, and deep WMH regions compared to women. Additionally, the risk factors for WMH development and progression differ between genders. In men, a history of hypertension was identified as the strongest contributor to WMH burden, whereas in women, a vascular composite was the most significant contributor. (Morrison et al., 2024).

Higher education is a widely recognized indicator of CR, and individuals with higher levels of education tend to have greater cognitive reserve. Research has demonstrated that higher education levels can mitigate the adverse impact of WMH on cognitive function and depressive symptoms. People with more education tend to experience fewer depressive symptoms and less cognitive decline, even as WMH levels rise (Lin et al., 2020).

The connection between education and WMH is still not fully understood. Nonetheless, increased education levels are usually linked to higher gray matter volume, cortical thickness, and surface areas of certain sub-regions of the amygdala and hippocampus (Seyedsalehi et al., 2023). Also, a higher educational level could improve efficiency in utilizing brain networks or alternative cognitive strategies to compensate for the damage caused by WMHs (Chen et al., 2019). Cognitive reserve allows individuals to optimize brain function by recruiting alternative networks or by using existing networks more effectively, which may explain why individuals with higher education are less affected by WMH burden (Stern, 2012). Therefore, it is suggested that higher education might enhance the brain’s resilience against WMH and other neuropathological insults.

Nevertheless, after evaluating the main effect of education in our statistical model, we determined it to be non-significant. This suggest that education alone may not directly reduce WMH burden, but rather it plays a crucial role in mitigating the effects of WMHs on cognitive decline when considered alongside other factors, such as the progression from HC to MCI.

An interesting finding from our statistical model was the significant interaction effect between education and group, indicating that the relationship between education and WMH burden varies depending on the group. Upon analyzing each group separately, we observed a different pattern between years of education and WMH load. Specifically, individuals who transitioned to MCI showed a decrease in WMH burden with higher education levels, whereas this trend was not seen in the HC remain group. This finding aligns with previous research suggesting that individuals with higher levels of education, which serves as a measure of cognitive reserve, may experience less cognitive decline even in the presence of WMHs (Dufouil et al., 2003; Murray et al., 2012). Thus, cognitive reserve may buffer against the negative effects of brain pathology, such as WMHs, by modulating how these pathologies affect cognitive outcomes (Zhong et al., 2023). Furthermore, cognitive reserve could mitigate the impact of brain pathology, thereby delaying or reducing the severity of cognitive decline in aging populations (Stern, 2002).

### Strengths and Limitations

The added value of our study lies in several key aspects that differentiate it from these earlier works. First, our study employs a longitudinal design focusing specifically on the transition from healthy cognitive status HC to MCI over a seven-year period. This long-term perspective allowed us to provide insights into how cognitive reserve, as measured by education, influences the progression of cognitive decline—an area not as thoroughly explored in previous research. The study’s limitations include the reliance on educational attainment as a proxy for cognitive reserve, which may not fully capture the multidimensional nature of cognitive reserve. Future research should explore other markers of cognitive reserve, such as occupational complexity or engagement in cognitively stimulating activities, to provide a more comprehensive understanding of how cognitive reserve interacts with brain pathology.

## 5- Conclusion

This study focused on the transition from cognitively healthy to MCI. Our results indicate that education, as a measure of cognitive reserve, plays a protective role in reducing the negative impact of WMH accumulation in older adults at risk of cognitive decline. These findings highlight the significance of educational attainment in promoting cognitive health and delaying the onset of cognitive impairment. Further research is needed to investigate the mechanisms underlying this protective effect and to develop strategies for enhancing cognitive reserve through educational and other cognitive enrichment activities.

## Data Availability

Data used in the preparation of this article were obtained from the Alzheimers Disease Neuroimaging Initiative (ADNI) database (adni.loni.usc.edu).

## Data Availability

Data used in the preparation of this article were obtained from the Alzheimer’s Disease Neuroimaging Initiative (ADNI) database (adni.loni.usc.edu). The ADNI was launched in 2003 as a public-private partnership, led by Principal Investigator Michael W. Weiner, MD. The primary goal of ADNI has been to test whether serial magnetic resonance imaging (MRI), positron emission tomography (PET), other biological markers, and clinical and neuropsychological assessment can be combined to measure the progression of mild cognitive impairment (MCI) and early Alzheimer’s disease (AD).

## Acknowledgments

Data collection and sharing for this project was funded by the Alzheimer’s Disease Neuroimaging Initiative (ADNI) (National Institutes of Health Grant U01 AG024904) and DOD ADNI (Department of Defense award number W81XWH-12-2-0012). ADNI is funded by the National Institute on Aging, the National Institute of Biomedical Imaging and Bioengineering, and through generous contributions from the following: AbbVie, Alzheimer’s Association; Alzheimer’s Drug Discovery Foundation; Araclon Biotech; BioClinica, Inc.; Biogen; Bristol-Myers Squibb Company; CereSpir, Inc.; Cogstate; Eisai Inc.; Elan Pharmaceuticals, Inc.; Eli Lilly and Company; EuroImmun; F. Hoffmann-La Roche Ltd and its affiliated company Genentech, Inc.; Fujirebio; GE Healthcare; IXICO Ltd.; Janssen Alzheimer Immunotherapy Research & Development, LLC.; Johnson & Johnson Pharmaceutical Research & Development LLC.; Lumosity; Lundbeck; Merck & Co., Inc.; Meso Scale Diagnostics, LLC.; NeuroRx Research; Neurotrack Technologies; Novartis Pharmaceuticals Corporation; Pfizer Inc.; Piramal Imaging; Servier; Takeda Pharmaceutical Company; and Transition Therapeutics. The Canadian Institutes of Health Research is providing funds to support ADNI clinical sites in Canada. Private sector contributions are facilitated by the Foundation for the National Institutes of Health (www.fnih.org). The grantee organization is the Northern California Institute for Research and Education, and the study is coordinated by the Alzheimer’s Therapeutic Research Institute at the University of Southern California. ADNI data are disseminated by the Laboratory for Neuro Imaging at the University of Southern California.

## Author Contributions

**OE:** Conceived the project, designed the study, conducted statistical analysis, interpreted results, and was responsible for writing and editing.

**IB:** Sorted data, created visualizations, and contributed to writing and editing.

## Conflicts of Interest

The authors have no conflicts of interest to declare.

## Notes

### Competing Interest Statement

The authors have declared no competing interest.

### Funding Statement

This study did not receive any funding

